# Economic Evaluation of Mindfulness-Oriented Recovery Enhancement for the Treatment of Opioid Misuse

**DOI:** 10.64898/2026.02.23.26346912

**Authors:** Fernando A. Wilson, Eric L. Garland

## Abstract

**OBJECTIVE:** Opioid misuse exacts a tremendous toll on society. Mindfulness-Oriented Recovery Enhancement (MORE) is an efficacious treatment for opioid misuse. Yet, the cost-effectiveness of this intervention remains unknown.

**METHODS:** Cost-effectiveness and cost-benefit analyses of a randomized clinical trial with enrollment of 250 adults with chronic pain prescribed long-term opioid therapy who were misusing opioids. Participants were randomized to MORE (training in mindfulness, reappraisal, and savoring positive experiences) or supportive group psychotherapy across 8 weekly 2-hour groups. Incremental cost-effectiveness ratios (ICER) and benefit-to-cost ratios (BCRs) were computed using the primary outcome of opioid misuse at 9-month follow-up, as assessed by a composite measure based on self-report, clinical interview, and urine screen.

**RESULTS:** 250 randomized patients (64.0% female) had an average age of 51.8 years (SD=11.9), were mostly taking oxycodone or hydrocodone (69%), and had mean morphine equivalent opioid dose of 101.0 (IQR=74) mg. At 9-mo. follow-up, the difference in the probability of having a positive Drug Misuse Index (DMI) rating was 0.24 (0.54 for MORE participants vs. 0.78 for controls). The ICER of MORE relative to supportive psychotherapy was $116.3 per averted case of opioid misuse, $8.9 per life-year, and $8.0 per quality-adjusted life-year. MORE is cost-saving vs. supportive psychotherapy after adjusting for healthcare costs. Excluding all benefits associated with averting fatal overdoses results in a BCR of 84.2.

**CONCLUSIONS:** Given MORE’s cost-effectiveness, private and public payers should consider disseminating this evidence-based therapy broadly across the nation to reduce mortality and morbidity associated with the ongoing opioid crisis.

*HIGHLIGHTS:*

- Mindfulness-Oriented Recovery Enhancement (MORE) substantially reduced opioid misuse among adults with chronic pain on long-term opioid therapy.
- MORE was highly cost-effective vs. supportive psychotherapy, costing $116 per averted opioid misuse case, and MORE was cost saving when accounting for healthcare costs associated with opioid misuse.
- Findings suggest wide dissemination of this evidence-based treatment could yield major healthcare and other economic benefits in addressing the opioid crisis.

## Introduction

Increased prescribing of long-term opioid therapy (LTOT) for chronic pain was a key vector for the first wave of the opioid crisis,^1^ a crisis that has exacted a tremendous toll on society. Approximately one-quarter of individuals receiving LTOT misuse opioids, which increases risk for opioid overdose and development of opioid use disorder (OUD).^2–4^ Escalation from LTOT to opioid misuse, OUD, and overdose raises numerous economic, health, and social costs. Opioid misuse and OUD are associated with increased inpatient stays and emergency department visits.^5,6^ OUD significantly increases the odds of homelessness.^7^ Moreover, the progression from prescription opioid use to opioid misuse and OUD is associated with increased rates of criminal justice system involvement.^8^ OUD also results in increased risk of death even while under the care of a physician; people with OUD are 1.6 times more likely to die during hospitalization than the general population.^9^ OUD has also been associated with an approximate 15-year loss in life expectancy for 18 year old individuals compared to the general population.^10^ Finally, opioid misuse and OUD have severe impacts on long-term economic productivity,^11^ with untreated OUD resulting in high productivity costs, with the average worker with OUD losing over 14 days of work per year.^12^

Therefore, interventions that reduce opioid misuse and prevent the progression to OUD can result in societal impact and cost savings from reductions in costly healthcare utilization and substance use treatment, fewer interactions with the criminal justice system, improved long-term economic productivity, and improvements in the individual’s quality of life. Recently, we reported results from a full-scale randomized clinical trial (RCT)^13^ of Mindfulness-Oriented Recovery Enhancement (MORE), a novel behavioral intervention that combines mindfulness training, cognitive-behavioral therapy (CBT), and principles from positive psychology into a treatment designed to simultaneously address addictive behavior, emotional distress, and chronic pain by normalizing brain reward function.^14^ The target population for MORE is patients with chronic pain with co-occurring prescription opioid misuse or OUD. MORE is an 8-session, manualized group treatment for people with chronic pain and co-occurring prescription opioid misuse or OUD. MORE integrates training in mindfulness skills to alleviate pain and craving and strengthen self-regulation over drug use, reappraisal skills to regulate negative emotions and cultivate meaning in life, and savoring skills to boost positive emotions and amplify natural reward responsiveness. In this trial (N=250), the overall odds ratio for reduction in opioid misuse across the entire 9-month follow-up period in the MORE group compared with a supportive psychotherapy control condition was 2.06, with a 2.94 odds ratio at the most distal 9-month follow-up endpoint.^13^ In addition, MORE was associated with significant reductions in chronic pain symptoms and opioid dosing through the 9-month follow-up. Here, we leverage outcome and cost data from this trial to conduct an economic evaluation of the MORE intervention.

## Methods

The cost-effectiveness analysis is based on data reported in the RCT of MORE vs. supportive group psychotherapy by Garland et al.^13^ This trial consisted of 250 patients (129 patients in MORE and 121 patients in the control group) recruited from 2016 to 2020 who were being treated in primary care clinics located in Utah. Patients were included if they were receiving long-term, daily opioid analgesic therapy for chronic pain (_≥_90 days) and were misusing opioids based on the Current Opioid Misuse Measure (COMM) (_≥_9 points). Patients receiving cancer treatment or those with suicidal behavior, psychosis, or severe non-opioid substance use disorder were excluded, as were those who had completed another mindfulness-based intervention in the past. Of the randomized sample, 64% were women, with a mean age of 51.8 years (SD=11.9). On average, participants reported pain for a mean of 14.7 years (range 1 to 60 years); the most common pain condition was low back pain (68%), but 76% of participants reported _≥_2 or more pain conditions. The mean pain severity score on the Brief Pain Inventory (BPI) was 5.5 (SD=1.5). The majority of participants (69%) were prescribed oxycodone or hydrocodone; the mean morphine equivalent daily opioid dose was 101.0 mg (SD=266.3 mg, IQR=74 mg).

MORE is an 8-session, manualized group treatment^15^ that integrates training in mindfulness skills to alleviate pain and craving and strengthen self-regulation over drug use, reappraisal skills to regulate negative emotions and cultivate meaning in life, and savoring skills to boost positive emotions and amplify natural reward responsiveness. The 8-session supportive psychotherapy group control condition involved Rogerian, client-centered therapy in which the clinician provided empathic responding and elicited emotional disclosure about topics related to chronic pain and opioid use.

Intervention and control costs include therapist compensation to deliver MORE and supportive psychotherapy, training costs for therapists to implement the MORE therapy, and supervision of the therapists to ensure fidelity. Therapy costs consist of eight 2-hour weekly sessions totaling 320 therapy hours for each trial arm, totaling $2,500 per patient cohort (consisting of 6-7 patients per cohort). Therapy costs were based on the actual gross costs from the underlying NIH study by Garland and colleagues which utilized a medical center perspective.^13^ In addition, included in the intervention costs, training in MORE entails an intensive, 13-hour training workshop that includes didactic and experiential instruction in the delivery of mindfulness, reappraisal, and savoring skills to address addiction, emotional distress, and chronic pain. Training also involves regular supervision guided by fidelity monitoring of session audio recordings. In addition, total supervision hours consisted of 32 hours for each arm. The net cost difference between the two trial arms is the cost of MORE training, or $900 per therapist. As detailed in the MORE study, participants who missed interim follow-up visits were allowed to re-enter at subsequent assessments, and no missing data were imputed. Primary analyses used intention-to-treat full information maximum likelihood estimation under a missing-at-random assumption. Additional sensitivity analyses were undertaken, including incorporating auxiliary covariates (e.g., durations of opioid treatment), not-missing-at-random selection models, and a pattern mixture control group switching modeling approach, in order to assess robustness of findings to alternative missingness mechanisms. However, the findings did not substantively change across approaches. For more information, refer to Supplemental Content provided by Garland et al(2022).^13^

We utilized the rates of opioid misuse at 9-months follow-up reported in Garland et al(2022). This was the longest-term (9-month) follow-up period in the trial, and we believed that using the longest-term follow-up period was most appropriate for our analysis. However, as reported below, using the findings at 3- and 6-months, did not substantially alter our analysis and conclusions. The pre-specified opioid misuse outcome was assessed with a validated composite measure - the Drug Misuse Index (DMI).16 The DMI uses three levels of data to characterize opioid misuse: 1) self-reports on the Current Opioid Misuse Measure (COMM); 2) clinical assessment of opioid misuse on the Addiction Behaviors Checklist (ABC)^17^ as rated by clinical staff (i.e., nurses, social workers, and psychologists) who were blinded to treatment assignment; and 3) urine toxicology screens. On the DMI, COMM scores _≥_9 and ABC scores _≥_2 were considered positive for opioid misuse. The urine screen was scored as positive when subjects tested positive for illicit drugs or non-prescribed opioids. Patients with positive COMM scores were given a positive DMI. If COMM scores did not meet the cutoff for opioid misuse (COMM scores<9), then positive ratings on both the ABC and urine screen were required for a positive DMI. This scoring approach was used because urine screens can be inaccurate due to false positives and variable drug metabolites, and clinician ratings may suffer from poor interrater reliability. Otherwise, subjects were given a negative DMI. Multiple other studies have employed this DMI scoring method.^16,18–21^ Data from the 2021 National Survey of Drug Use and Health (NSDUH) were analyzed to determine the rate of opioid misuse and opioid use disorder in Utah around the time of the completion of the trial (January 2020). The NSDUH is a nationally and state representative survey of the civilian non-institutionalized population aged 12 and older, conducted annually and consisting of both in-person and web-based interviews. In NSDUH, opioid misuse is defined as use of a prescription opioid in any way not directed by a doctor, and OUD is measured according to DSM-5 criteria, classifying respondents who meet this diagnostic threshold within in the past year as diagnosed with OUD. The NSDUH estimates were utilized to estimate the likelihood of transition from opioid misuse to OUD in Utah.

The cost-effectiveness analyses consisted of multiple approaches. First, we estimated incremental cost-effectiveness ratios (ICER) to determine the incremental per-patient cost of MORE vs. control relative to the decrease in number of opioid misuse cases. Per-patient cost of MORE consists of training cost per therapist ($900 in 2025), number of hours (therapy and supervision) to deliver the MORE intervention, and the US distribution of hourly wage compensation for mental health and substance use social workers and for medical/health services managers for supervision. Second, we utilized prior research on healthcare cost savings from averting opioid use disorder, and subtracted these estimates from the intervention cost of MORE. Finally, we conducted sensitivity analyses based on three cost scenarios: (1) Baseline Cost: per-patient cost of delivering supportive psychotherapy to controls is zero; (2) Intermediate Cost: per-patient staffing costs of MORE and delivering supportive psychotherapy are equal, and thus the incremental cost of MORE is the average therapist training cost per patient; and (3) High Cost: per-patient staffing costs of MORE are at the 90% percentile of the wage distribution and the cost of delivering supportive psychotherapy is zero. Because the trial occurred within a 9-month period, the ICERs were not discounted.

In addition to CEA, we utilized prior research on long-term or life-time monetary cost savings from decreasing OUD in order to conduct a cost-benefit analysis, using benefit-to-cost ratios (BCR), of MORE vs. controls. The BCR analysis was based on the ratio of the monetary impact of MORE (vs. supportive psychotherapy) in decreasing opioid misuse, thus decreasing risk of misuse leading to OUD and fatal overdose (using NSDUH and fatality rate data), and resulting in consequent monetary cost savings from averting OUD, divided by the difference in average cost of MORE per patient based on compensation provided to staff during the clinical trial relative to supportive psychotherapy. Thus, the BCR provides a measure of net economic value generated for each dollar invested in MORE vs. supportive psychotherapy. Fatality rate data associated with opioid use were reported by Bahji et al.^22^ Bahji et al(2020) is a systematic review and meta-analysis of mortality rates for individuals with OUD, which included 15 studies conducted in the US. For these studies, the meta-analysis reported a mortality rate of 17.1 per 1,000 person-years. Average estimated economic benefit per patient utilized data provided by Florence et al.(2021)^11^ on the economic burden of OUD and fatal opioid overdose in the US. This study utilized multiple databases in order to estimate healthcare, criminal justice, and lost productivity costs associated with OUD cases. For healthcare costs, Florence and colleagues used the IBM MarketScan Research Database of commercial, Medicaid and Medicare covered individuals in order to estimate healthcare expenditures attributable to treatment of OUD. Criminal justice costs used data from the Justice Expenditure and Employment Extracts, and included police protection, legal and adjudication, correctional facilities, and lost property costs. Lost productivity costs were estimated based on premature death due to overdose, decreased productive hours, and incarceration. These cost data were based on analyses of the Web-based Injury Statistics Query and Reporting System maintained by the Centers for Disease Control and Prevention. In addition, the study used data and methodologies from the NSDUH, National Drug Intelligence Center, Drug Enforcement Administration, and prior published studies.

Microsimulation analysis was undertaken to determine the distribution of BCRs by age and perspective when scaling MORE to 1,000 patients residing in Utah. The analysis was conducted from healthcare, taxpayer and societal perspectives using cost data from Murphy(2020).^23^ This study analyzed data from NSDUH and the Centers for Disease Control and Prevention’s WONDER database to estimate economic value of averting OUD by year of age and stratified by healthcare, taxpayer, and societal perspectives. A discount rate was utilized by Murphy(2020) to discount future costs. The microsimulation approach models individual-level variation in the risk of OUD by generating random draws from a normal distribution based on the prevalence of opioid misuse and OUD in Utah. Sensitivity analyses were conducted using minimum and maximum range estimates for healthcare, taxpayer and societal costs reported in Murphy(2020).

Our study conforms to the Cost-Effectiveness Analysis Alongside Clinical Trials II and the Consolidated Health Economic Evaluation Reporting Standards (CHEERS)^24^ checklist. Cost estimates were adjusted for inflation to 2023 dollars using the Consumer Price Index.

## Results

Model parameter values for the cost-effectiveness analysis are provided in Table 1. These data are based on estimates provided by Garland et al.^13^ and via correspondence with the authors. Our analysis is based on 20 cohorts in total for each arm. Therapy hours and compensation are identical. However, the training cost of MORE was $900 per therapist, and thus the average cost per patient of MORE is $406 versus $388 for supportive psychotherapy. At 9-months follow-up, 54% of MORE participants and 78% of participants in supportive psychotherapy had opioid misuse based on the DMI—a between-groups difference of 24 percentage points (odds ratio at 9-months=2.94, p<0.01, as previously reported). Data from the 2021 National Survey of Drug Use and Health (NSDUH) indicate that 3.3% of Utahns misuse opioids and 1.7% have opioid use disorder. The fatality rate from OUD is 17.10 per 1,000 person-years.^22^

**Table 1.** Model parameter values for cost-effectiveness analysis.

| Characteristic | MORE | Supportive Psychotherapy | Source |
| --- | --- | --- | --- |
| **Program-level** |  |  |  |
| Total patients, # | 129 | 129 | Garland et al.<sup>12</sup> |
| Patient cohorts, # | 20 | 20 | Garland et al.<sup>12</sup> |
| Therapy time, hours* | 16 | 16 | Garland et al.<sup>12</sup> |
| Total therapy hours | 320 | 320 | Garland et al.<sup>12</sup> |
| Supervision, hours | 8 | 8 | Garland et al.<sup>12</sup> |
| Number of therapists | 4 | 4 | Garland et al.<sup>12</sup> |
| Total supervision hours | 32 | 32 | Garland et al.<sup>12</sup> |
| **Cost estimates** |  |  |  |
| MH/SU social worker, median wage | 28.88 | 28.88 | BLS OEWSQS |
| Medical services manager, median wage | 56.71 | 56.71 | BLS OEWSQS |
| Therapist compensation per cohort, \$ | 2,500 | 2,500 | Garland et al.<sup>12</sup> |
| MORE training cost per therapist, \$ | 900 | n/a | Garland et al.<sup>12</sup> |
| Probability of opioid misuse at 9-months | 0.54 | 0.78 | Garland et al.<sup>12</sup> |
| **Transition probabilities** |  |  |  |
| Prevalence of misuse | 0.033 |  | Authors' analysis of 2021 NSDUH |
| Prevalence of OUD | 0.017 |  | Authors' analysis of 2021 NSDUH |
| Opioid-related fatality rate per 1,000 person-years | 17.1 |  | Bahji et al.<sup>21</sup> |
| 10-year fatality rate | 0.16 |  | Bahji et al.<sup>21</sup> |

MORE, Mindfulness-Oriented Recovery Enhancement; OUD, Opioid Use Disorder; BLS, Bureau of Labor Statistics; OEWSQS, Occupational Employment and Wage Statistics Query System; NSDUH, National Survey of Drug Use and Health
\* Conducted in eight 2-hour sessions.

### Incremental Cost-Effectiveness Analysis

In Table 2, incremental effectiveness utilizes the 9-month decrease in probability of opioid misuse of MORE vs. supportive psychotherapy (0.24, or 0.54 vs. 0.78, respectively). Incremental cost is an average of $27.9 per person, resulting in an ICER of $116.3 per averted misuse case. Using outcomes from 3-, 6-, or 9-months follow-up resulted in the ICERs ranging from $116.30 to $232.50. To estimate number of life-years saved, we utilize the estimated 15-year loss in life expectancy associated with OUD provided by Lewer and colleagues(2020).^10^ Using this estimate, the likelihood of transitioning from misuse to OUD (described previously), and Social Security Administration (SSA) actuarial life-tables^25^, which provided life expectancies for the general US population by year of age, we calculated expected number of life-years per person based on either developing OUD (resulting in only 15 life-years) or not developing OUD (estimated 31.8 life-years for MORE (average age was 51) and 30.08 life-years for the control sample (average age was 53), using the SSA data). Using this approach, expected life-years for the supportive psychotherapy sample was estimated to be 24.0 life-years, and 27.1 life-years for the MORE sample, or a difference of 3.1 life-years. Based on the incremental cost difference of $27.9, this suggests an ICER of $8.9 per life-year. A separate regression analysis of quality of life estimates suggests that individuals with OUD and moderate symptoms lose 0.183 in health state utility value.^26^ Thus, we adjusted estimated life-years for OUD by 0.817 (1 – 0.183), and used the same process as above to estimate the incremental increase in quality-adjusted life-years (QALY) between MORE and the control group. This resulted in a difference of 3.5 QALYs, and an ICER of $8.0 per QALY (Table 2). Using typical decision rules of $50,000, $100,000, or $150,000 per QALY, this suggests that MORE is highly cost-effective.^27,28^

**Table 2.**
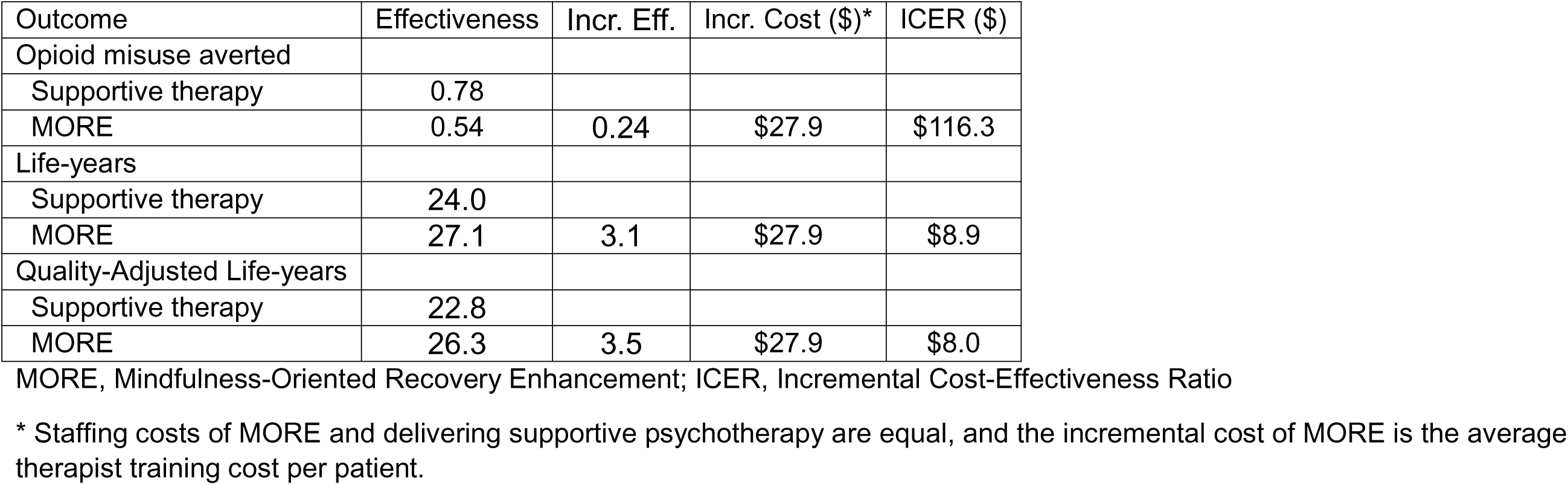
Incremental cost-effectiveness analysis of MORE vs. supportive psychotherapy.

| Outcome | Effectiveness | Incr. Eff. | Incr. Cost (\$)* | ICER (\$) |
| --- | --- | --- | --- | --- |
| Opioid misuse averted |  |  |  |  |
| Supportive therapy | 0.78 |  |  |  |
| MORE | 0.54 | 0.24 | \$27.9 | \$116.3 |
| Life-years |  |  |  |  |
| Supportive therapy | 24.0 |  |  |  |
| MORE | 27.1 | 3.1 | \$27.9 | \$8.9 |
| Quality-Adjusted Life-years |  |  |  |  |
| Supportive therapy | 22.8 |  |  |  |
| MORE | 26.3 | 3.5 | \$27.9 | \$8.0 |

MORE, Mindfulness-Oriented Recovery Enhancement; ICER, Incremental Cost-Effectiveness Ratio
\* Staffing costs of MORE and delivering supportive psychotherapy are equal, and the incremental cost of MORE is the average therapist training cost per patient.

The analysis in Table 2 does not account for healthcare or other economic cost savings from averting opioid misuse. Table A1 in Supplemental Materials re-estimates ICERs adjusting for healthcare-related cost savings from averting one case of opioid misuse, using cost savings estimates reported in Murphy(2020).^23^ The scenarios for the sensitivity analyses are based on the wage distribution of clinical social workers and average cost of supportive psychotherapy. Table A2 presents sensitivity analyses of MORE vs. supportive psychotherapy under three program cost scenarios. For example, the ICER for opioid misuse averted ranges from $473.4 to $742.6 under the highest cost scenario. MORE therapists may include a range of provider types including clinical psychologists who have 60% higher median wage compensation than social workers. Using wages at the 90% percentile for clinical psychologists and for managers/supervisors, this results in an ICER of $1,071 (results not shown).

### Benefit-to-Cost Ratio Analysis

In addition to the ICER analysis, cost-benefit analyses were undertaken using cost data provided by the clinical trial on staffing. Using data on OUD and opioid overdose from Florence et al.(2021)^11^ and Luo et al.(2021)^29^, Table A3 presents the distribution of costs per opioid overdose including expenditures from non-fatal healthcare utilization, substance use treatment, interactions with the criminal justice system including police enforcement, prosecution and incarceration, decreased labor market productivity such as lost workhours and costs associated with a fatal overdose including the value of lost life-years. Based on the prevalence of opioid misuse and OUD in the State of Utah and the National Survey of Drug Use and Health, we estimated that more than half (52.7%) of those who misuse opioids will progress to OUD. Using these estimates and the efficacy of MORE in decreasing likelihood of opioid misuse, this results in a lifetime economic impact totaling $324,489 per participant, primarily due to the value of a statistical life and mortality reduction.^10,29^ This suggests that the benefit-to-cost ratio of MORE relative to supportive psychotherapy is 781, and, excluding all benefits associated with averting fatal overdoses results in a BCR of 84.2 (results not shown).

There are significant cost savings associated with MORE across third party payers. For example, providing MORE to privately insured individuals is predicted to result in $959 in cost savings per participant (Figure A1). For Medicaid, these cost savings are $828 per participant. Monte Carlo microsimulation was utilized to predict the costs and benefits of expanding MORE to a population of 1,000 participants ranging in age from 18 to 71. Using supplemental data provided by Murphy(2020)^23^ on the cost of OUD from the healthcare system perspective, we estimated the predicted BCR by year of age (Figure 1). BCRs associated with healthcare cost savings from MORE are highest for younger age groups, ranging from 72.9 (low estimate) to 213.6 (high estimate) for ages 18 to 23. Thus, although MORE is likely to result in cost savings relative to supportive psychotherapy across all age groups, it has highest impact for younger participants.

**Figure 1.**
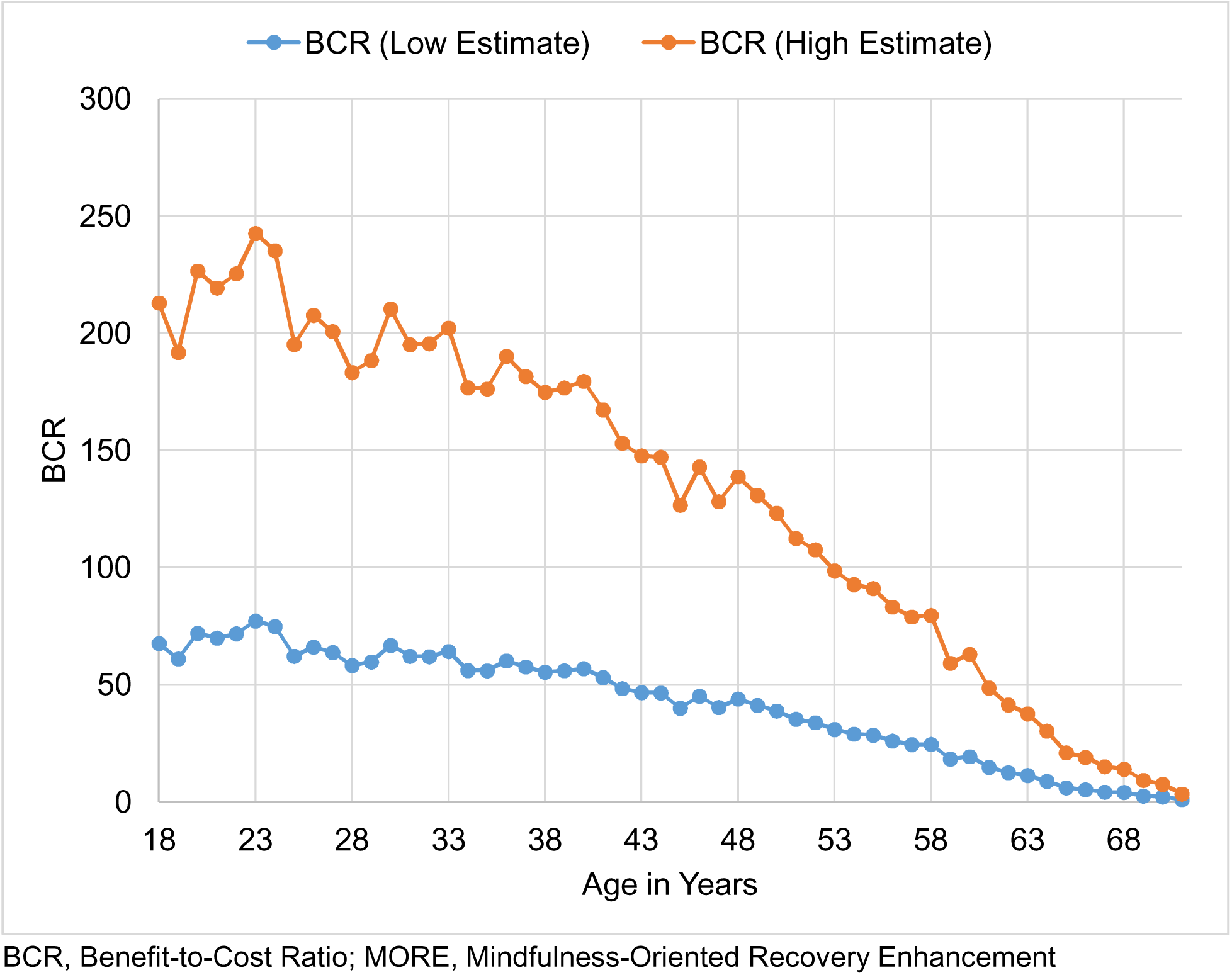
Predicted benefit-to-cost ratios (BCR) from averted healthcare costs by participant age from participation in MORE therapy relative to supportive psychotherapy

## Discussion

Results from this analysis demonstrate substantial cost-effectiveness in favor of MORE, an evidence-based intervention for the treatment of opioid misuse among people with chronic pain.^30,31^ At 9-months follow-up, the MORE intervention decreased the likelihood of opioid misuse by over 30% relative to the supportive psychotherapy control condition. Given the risk and broad and substantial adverse impact of OUD, this large effect size reduction in opioid misuse among patients treated with MORE has a wide range of potential economic impacts stemming from reduced health care costs, decreased substance use disorder treatment services and criminal justice involvement, and increased labor productivity and quality of life. Although our analysis examines the primary outcome of opioid misuse, the parent trial for the analysis reported additional significant outcomes for the MORE intervention relative to supportive psychotherapy.^13^ At 9-months follow-up, mean Brief Pain Inventory severity scores, Depression Anxiety Stress Scale scores, and morphine equivalent daily opioid dose were also significantly lower for MORE vs. the control group. As such, our cost-effectiveness analyses may be conservative, and the improvements in pain, depression and anxiety are likely to lead to substantial benefits to patients’ quality of life and to reduced likelihood of relapse.^32^

The ICER analysis suggests that MORE is highly cost-effective resulting in an ICER of $116 per one case decrease in opioid misuse. Incorporating estimated healthcare costs savings from averting opioid misuse resulted in MORE being cost-saving relative to supportive psychotherapy. Furthermore, the BCRs observed for MORE exceed those observed for other interventions for people exhibiting addictive opioid use behaviors. Cost-effectiveness research of interventions to treat OUD has identified BCRs of 1.8 for comprehensive case management, 2.0 to 4.8 for residential treatment, 5.1 for intensive outpatient, and 0.9 to 37.7 for medication assisted treatment.^33–38^ The substantial cost-effectiveness MORE observed in the present study may be due to several reasons. First, the efficacy of MORE is high; MORE nearly tripled the effect of the active supportive psychotherapy control condition at the 9-month follow-up (OR=2.94), with an absolute reduction in the rate of opioid misuse of 45%. Recent meta-analytic evidence indicates that 29.6% of chronic pain patients treated with opioid analgesics develop signs and symptoms of dependence and OUD.^4^ However, all participants in Garland et al.^13^ were misusing opioids at baseline, and thus the likelihood of these individuals going on to develop full blown OUD was high. Because this is based on a single year of data, this rate may also be significantly underestimated for individuals misusing opioids over time. In addition, MORE may not only prevent the transition from opioid misuse to OUD; MORE has demonstrated efficacy as a treatment for OUD. In that regard, a more recent trial found that MORE reduced relapse rates by 42% and dropout from addictions treatment by 59%^39^ among patients receiving methadone treatment for OUD. In addition, the training costs associated with MORE were low at the time of this study. Finally, relative to its cost, MORE is predicted to generate substantial economic impact across a range of healthcare, criminal justice, labor market and other societal outcomes.

Prior research estimated healthcare cost savings of $1,277 (inflation-adjusted) for a mindfulness-based stress reduction vs. usual care for adults with chronic low back pain.^40^ Aside from healthcare costs, criminal justice-related expenditures (police, legal, corrections, lost property and productivity) average over $13,000 per OUD case. Furthermore, there are also substantial impacts on lost labor productivity, quality of life and premature mortality.^11,22,29^ A substantial proportion of MORE’s estimated benefit is in decreased risk of opioid overdose and thus averted premature mortality among patients misusing opioids and at high-risk of OUD, resulting in a lifetime economic impact of over $324 thousand.^11,23^

Results from this study should be interpreted in the context of certain limitations. Healthcare utilization or expenditure data were not reported for patients in Garland et al.(2022). Prior studies by Florence et al.(2021) and Luo et al.(2021) provide detailed, stratified expenditure data for healthcare and non-healthcare settings associated with opioid use disorder and overdose. However, data from prior literature may not represent the healthcare utilization of people using the MORE study or reflect healthcare activity of people living in Utah. Similarly, long-term follow-up data on the progression of patients from misuse to diagnosed OUD were not available. Finally, the analysis is based on data for the State of Utah which was the location of the MORE intervention in this trial. New implementation science studies are now examining the scaling of MORE to other geographic locations (e.g., California, New Jersey) and care settings (e.g., OUD treatment settings, orthopedic surgery).

## Conclusions

Our economic evaluation suggests that MORE is a highly cost-effective treatment for opioid misuse. Given MORE’s clear cost-effectiveness, private and public payers should consider disseminating this evidence-based therapy broadly across the nation (potentially via opioid litigation settlement monies now reaching the states) to reduce mortality and morbidity associated with the ongoing opioid crisis.

## Data Availability

All data produced in the present study are available upon reasonable request to the authors

## Acknowledgments

None

## References

1. Compton WM, Jones CM. Epidemiology of the US opioid crisis: the importance of the vector. Ann N Y Acad Sci. 2019;1451(1):130–143.

2. Vowles KE, McEntee ML, Julnes PS, Frohe T, Ney JP, van der Goes DN. Rates of opioid misuse, abuse, and addiction in chronic pain: a systematic review and data synthesis. Pain. 2015;156(4):569–576.

3. Chou R, Turner JA, Devine EB, et al. The Effectiveness and Risks of Long-Term Opioid Therapy for Chronic Pain: A Systematic Review for a National Institutes of Health Pathways to Prevention Workshop. Ann Intern Med. 2015;162(4):276. doi:10.7326/M14-2559

4. Thomas KH, Dalili MN, Cheng HY, et al. Prevalence of problematic pharmaceutical opioid use in patients with chronic non-cancer pain: A systematic review and meta-analysis. Addiction. 2024;119(11):1904–1922. doi:10.1111/add.16616

5. Zhu H, Wu LT. National trends and characteristics of inpatient detoxification for drug use disorders in the United States. BMC Public Health. 2018;18(1):1073. doi:10.1186/s12889-018-5982-8

6. Weiss AJ, Elixhauser A, Barrett ML, Steiner CA, Bailey MK, O’Malley L. Opioid-related inpatient stays and emergency department visits by state, 2009–2014: statistical brief# 219. HCUP Stat Brief. Published online 2016. Accessed October 20, 2023. https://europepmc.org/books/n/hcupsb/sb219/?extid=21413206&src=med

7. Manhapra A, Stefanovics E, Rosenheck R. The association of opioid use disorder and homelessness nationally in the veterans health administration. Drug Alcohol Depend. 2021;223:108714.

8. Winkelman TN, Chang VW, Binswanger IA. Health, polysubstance use, and criminal justice involvement among adults with varying levels of opioid use. JAMA Netw Open. 2018;1(3):e180558–e180558.

9. Singh JA, Cleveland JD. National US time-trends in opioid use disorder hospitalizations and associated healthcare utilization and mortality. PLoS One. 2020;15(2):e0229174.

10. Lewer D, Jones NR, Hickman M, Nielsen S, Degenhardt L. Life expectancy of people who are dependent on opioids: A cohort study in New South Wales, Australia. J Psychiatr Res. 2020;130:435–440. doi:10.1016/j.jpsychires.2020.08.013

11. Florence C, Luo F, Rice K. The economic burden of opioid use disorder and fatal opioid overdose in the United States, 2017. Drug Alcohol Depend. 2021;218:108350.

12. Henke RM, Ellsworth D, Wier L, Snowdon J. Opioid use disorder and employee work presenteeism, absences, and health care costs. J Occup Environ Med. 2020;62(5):344–349.

13. Garland EL, Hanley AW, Nakamura Y, et al. Mindfulness-Oriented Recovery Enhancement vs supportive group therapy for co-occurring opioid misuse and chronic pain in primary care: A randomized clinical trial. JAMA Intern Med. 2022;182(4):407–417.

14. Garland EL, Hudak J, Hanley AW, Bernat E, Froeliger B. Positive Emotion Dysregulation in Opioid Use Disorder and Normalization by Mindfulness-Oriented Recovery Enhancement: A Secondary Analysis of a Randomized Clinical Trial. JAMA Psychiatry. Published online April 30, 2025. doi:10.1001/jamapsychiatry.2025.0569

15. Garland EL. Mindfulness-Oriented Recovery Enhancement: An Evidence-Based Treatment for Chronic Pain and Opioid Use. Guilford Press; 2024.

16. Jamison RN, Ross EL, Michna E, Chen LQ, Holcomb C, Wasan AD. Substance misuse treatment for high-risk chronic pain patients on opioid therapy: A randomized trial. PAIN. 2010;150(3):390–400. doi:10.1016/j.pain.2010.02.033

17. Wu SM, Compton P, Bolus R, et al. The addiction behaviors checklist: validation of a new clinician-based measure of inappropriate opioid use in chronic pain. J Pain Symptom Manage. 2006;32(4):342–351.

18. Wasan AD, Ross EL, Michna E, et al. Craving of Prescription Opioids in Patients With Chronic Pain: A Longitudinal Outcomes Trial. J Pain. Published online 2012.

19. Wasan AD, Butler SF, Budman SH, et al. Does report of craving opioid medication predict aberrant drug behavior among chronic pain patients? Clin J Pain. 2009;25:193–198.

20. Wasan AD, Michna E, Edwards RR, et al. Psychiatric Comorbidity Is Associated Prospectively with Diminished Opioid Analgesia and Increased Opioid Misuse in Patients with Chronic Low Back Pain: Anesthesiology. 2015;123(4):861–872. doi:10.1097/ALN.0000000000000768

21. Lawrence R, Mogford D, Colvin L. Systematic review to determine which validated measurement tools can be used to assess risk of problematic analgesic use in patients with chronic pain. BJA Br J Anaesth. 2017;119(6):1092–1109. doi:10.1093/bja/aex316

22. Bahji A, Cheng B, Gray S, Stuart H. Mortality Among People With Opioid Use Disorder: A Systematic Review and Meta-analysis. J Addict Med. 2020;14(4):e118. doi:10.1097/ADM.0000000000000606

23. Murphy SM. The cost of opioid use disorder and the value of aversion. Drug Alcohol Depend. 2020;217:108382.

24. Husereau D, Drummond M, Augustovski F, et al. Consolidated Health Economic Evaluation Reporting Standards 2022 (CHEERS 2022) statement: updated reporting guidance for health economic evaluations. MDM Policy Pract. 2022;7(1):23814683211061097. doi:10.1177/23814683211061097

25. Social Security Administration. Actuarial Life Table. 2022. Accessed February 22, 2026. https://www.ssa.gov/oact/STATS/table4c6.html

26. Patton T, Boehnke JR, Goyal R, et al. Analyzing quality of life among people with opioid use disorder from the National Institute on Drug Abuse Data Share initiative: implications for decision making. Qual Life Res Int J Qual Life Asp Treat Care Rehabil. 2024;33(10):2783–2796. doi:10.1007/s11136-024-03729-6

27. Neumann PJ, Cohen JT, Weinstein MC. Updating cost-effectiveness: the curious resilience of the $50,000-per-QALY threshold. N Engl J Med. 2014;371(9):796–97.

28. Grosse SD. Assessing cost-effectiveness in healthcare: history of the $50,000 per QALY threshold. Expert Rev Pharmacoecon Outcomes Res. 2008;8(2):165–78.

29. Luo F. State-level economic costs of opioid use disorder and fatal opioid overdose—United States, 2017. MMWR Morb Mortal Wkly Rep. 2021;70. Accessed May 28, 2025. https://www.cdc.gov/mmwr/volumes/70/wr/mm7015a1.htm

30. Garland EL. Mindfulness-Oriented Recovery Enhancement: Implementing an evidence-based intervention for chronic pain, opioid use, and opioid addiction in clinical settings. Br J Clin Pharmacol. 2024;90(12):3028–3035. doi:10.1111/bcp.16147

31. Parisi A, Roberts RL, Hanley AW, Garland EL. Mindfulness-Oriented Recovery Enhancement for Addictive Behavior, Psychiatric Distress, and Chronic Pain: A Multilevel Meta-Analysis of Randomized Controlled Trials. Mindfulness. 2022;13(10):2396–2412.

32. Leung J, Santo Jr T, Colledge-Frisby S, et al. Mood and anxiety symptoms in persons taking prescription opioids: a systematic review with meta-analyses of longitudinal studies. Pain Med. 2022;23(8):1442–1456.

33. Mauser E, Van Stelle KR, Moberg DP. The Economic Impact of Diverting Substance-Abusing Offenders into Treatment. Crime Delinquency. 1994;40(4):568–588. doi:10.1177/0011128794040004006

34. Gerstein DR, Johnson RA, Harwood HJ, Fountain D, Suter N, Malloy K. Evaluating recovery services: the California drug and alcohol treatment assessment (CalDATA) general report. Publ No ADP. Published online 1994:94-629.

35. Harwood HJ, Fountain D, Livermore G. The Economic Costs of Alcohol and Drug Abuse in the United States, 1992. US Government Printing Office; 1998. Accessed May 28, 2025. https://books.google.com/books?hl=en&lr=&id=2nVrAAAAMAAJ&oi=fnd&pg=PA11&dq=hj+harwood+drug+&ots=3ajLMv07qB&sig=avYPmxux_djAHDmCPjBul5_zQBc

36. McCollister KE, French MT. The economic cost of substance abuse treatment in criminal justice settings. Treat Drug Offenders Policies Issues. Published online 2002:22–37.

37. Koenig L, Siegel JM, Harwood H, et al. Economic benefits of substance abuse treatment: Findings from Cuyahoga County, Ohio. J Subst Abuse Treat. 2005;28(2):S41–S50.

38. Zarkin GA, Dunlap LJ, Hicks KA, Mamo D. Benefits and costs of methadone treatment: results from a lifetime simulation model. Health Econ. 2005;14(11):1133–1150. doi:10.1002/hec.999

39. Cooperman NA, Lu SE, Hanley AW, et al. Telehealth mindfulness-oriented recovery enhancement vs usual care in individuals with opioid use disorder and pain: a randomized clinical trial. JAMA Psychiatry. 2024;81(4):338–346.

40. Herman PM, Anderson ML, Sherman KJ, Balderson BH, Turner JA, Cherkin DC. Cost-effectiveness of Mindfulness-based Stress Reduction Versus Cognitive Behavioral Therapy or Usual Care Among Adults With Chronic Low Back Pain. Spine. 2017;42(20):1511. doi:10.1097/BRS.0000000000002344

