## Supplemental Material for "Economic Evaluation of Mindfulness-Oriented Recovery Enhancement for the Treatment of Opioid Misuse"

Fernando A. Wilson, PhD[1,2,3]

Eric L. Garland, PhD, LCSW[4,5]

[1] Population Health Sciences, University of Utah, Salt Lake City, UT, USA
[2] Department of Economics, University of Utah, Salt Lake City, UT, USA
[3] Matheson Center for Health Care Studies, University of Utah, Salt Lake City, UT, USA
[4] Department of Psychiatry, University of California San Diego, La Jolla, CA, USA
[5] Sanford Institute for Empathy and Compassion, University of California San Diego, La Jolla, CA, USA

**Supplemental Material**

Table A1. Incremental cost-effectiveness analysis of MORE vs. supportive psychotherapy adjusting for healthcare cost savings

| Outcome | Effectiveness | Incr. Eff. | Incr. Cost ($)* | ICER ($) |
|---|---|---|---|---|
| Opioid misuse averted* | | | | |
| Supportive therapy | 0.78 | | | |
| MORE | 0.54 | 0.24 | -$68,057 | CS |
| Life-years | | | | |
| Supportive therapy | 24.0 | | | |
| MORE | 27.1 | 3.1 | -$68,057 | CS |
| Quality-Adjusted Life-years | | | | |
| Supportive therapy | 22.8 | | | |
| MORE | 26.3 | 3.5 | -$68,057 | CS |

MORE, Mindfulness-Oriented Recovery Enhancement; ICER, Incremental Cost-Effectiveness Ratio; CS, Cost Saving

* Staffing costs of MORE and delivering supportive psychotherapy are equal, and the incremental cost of MORE is the average therapist training cost per patient. Intervention cost is net of average healthcare cost savings of averting opioid misuse using healthcare cost estimates from Murphy(2020).

Table A2. Sensitivity analyses in incremental cost-effectiveness of MORE vs. supportive psychotherapy of averting opioid misuse

| Opioid misuse averted | Incremental effectiveness | Incremental cost ($) | ICER ($) |
|---|---|---|---|
| (1) Intermediate cost | 0.24 | 113.6 | 473.4 |
| (2) High cost | 0.24 | 178.2 | 742.6 |
| Life-years gained | Incremental effectiveness | Incremental cost ($) | ICER ($) |
| (1) Intermediate cost | 3.1 | 113.6 | 36.2 |
| (2) High cost | 3.1 | 178.2 | 56.8 |
| QALYs gained | Incremental effectiveness | Incremental cost ($) | ICER ($) |
| (1) Intermediate cost | 3.5 | 113.6 | 32.6 |
| (2) High cost | 3.5 | 178.2 | 51.1 |

MORE, Mindfulness-Oriented Recovery Enhancement; ICER, Incremental Cost-Effectiveness Ratio

Baseline cost: Staffing costs of MORE and delivering supportive psychotherapy are equal, and the incremental cost of MORE is the average therapist training cost per patient.

Intermediate cost: Median wages for staff. Supportive psychotherapy costs are zero.

High cost: Wages for staff are at 90% percentile of wage distribution. Supportive psychotherapy costs are zero.

Table A3. Distribution of predicted costs per opioid overdose by source of expenditure (adapted from Florence et al(2021).

| Item | Cost, $ |
|---|---|
| Non-fatal costs | |
| Healthcare expenditures | |
| Private Insurance | 7,575 |
| Medicare | 1,861 |
| Medicaid | 6,542 |
| Champus/VA | 660 |
| Other | 481 |
| Uninsured | 1,263 |
| Substance use treatment | |
| Federal | 496 |
| State/local | 1,366 |
| Private | 214 |
| Criminal Justice | |
| Police protection | 3,645 |
| Legal and adjudication | 1,655 |
| Correctional facilities | 320 |
| Property loss | 204 |
| Lost production of incarcerated | 4,598 |
| Labor market productivity | |
| Decrease productivity | 13,785 |
| Value of reduced quality of life | 228,983 |

| | |
|---|---|
| Fatal costs | |
| Lost productivity | 1,803,939 |
| Healthcare | 6,828 |
| VSL | 12,624,396 |

VSL, Value of Statistical Life

* Inflation-adjusted to 2023 dollars using Consumer Price Index

Figure A1. Predicted healthcare cost savings per patient from MORE by insurance status from participation in MORE therapy relative to supportive psychotherapy

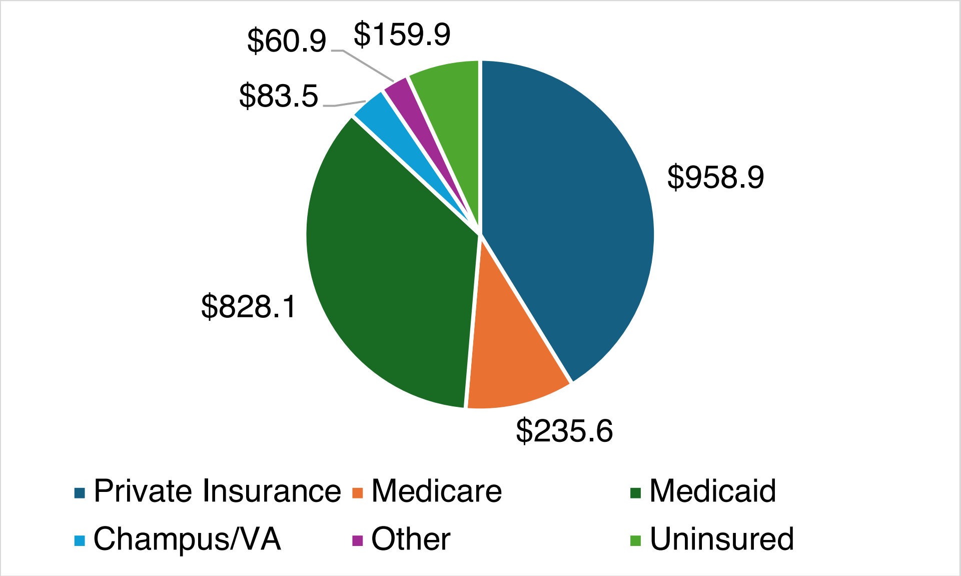

MORE, Mindfulness-Oriented Recovery Enhancement
